# Symptom Monitoring based on Digital Data Collection During Inpatient Treatment of Schizophrenia Spectrum Disorders – a Feasibility Study

**DOI:** 10.1101/2021.10.01.21264398

**Authors:** Julian Herpertz, Maike Frederike Richter, Carlotta Barkhau, Michael Storck, Rogério Blitz, Lavinia A. Steinmann, Janik Goltermann, Udo Dannlowski, Bernhard T. Baune, Julian Varghese, Martin Dugas, Rebekka Lencer, Nils Opel

**Author notes:** **Correspondence:** N. Opel, Department of Psychiatry, University of Münster, Albert-Schweitzer-Str. 11, 48149 Münster, Germany. This is to indicate that RL and NO contributed equally to the present work and should therefore both be considered senior author.

## Abstract

**Background:** Digital acquisition of risk factors and symptoms based on patients’ self-reports represents a promising, cost-efficient and increasingly prevalent approach for standardized data collection in psychiatric clinical routine. While the feasibility of digital data collection has been demonstrated across a range of psychiatric disorders, studies investigating digital data collection in schizophrenia spectrum disorder patients are scarce. Hence, up to now our knowledge about the acceptability and feasibility of digital data collection in patients with a schizophrenia spectrum disorder remains critically limited.

**Objective:** The objective of this study was to explore the acceptance towards and performance with digitally acquired assessments of risk and symptom profiles in patients with a schizophrenia spectrum disorder in comparison with patients with an affective disorder.

**Methods:** We investigated the acceptance, the required support and the data entry pace of patients during a longitudinal digital data collection system of risk and symptom profiles using self-reports on tablet computers throughout inpatient treatment in patients with a schizophrenia spectrum disorder. As a benchmark comparison, findings in patients with schizophrenia spectrum disorder were evaluated in direct comparison with findings in affective disorder patients. The influence of sociodemographic data and clinical characteristics on the assessment was explored. The study was performed at the Department of Psychiatry at the University of Münster between February 2020 and February 2021.

**Results:** Of 82 patients diagnosed with a schizophrenia spectrum disorder who were eligible for inclusion 59.8% (n=49) agreed to participate in the study of whom 54.2% (n=26) could enter data without any assistance. Inclusion rates, drop-out rates and subjective experience ratings did not differ between patients with a schizophrenia spectrum disorder and patients with an affective disorder. Out of all participating patients, 98% reported high satisfaction with the digital assessment. Patients with a schizophrenia spectrum disorder needed more support and more time for the assessment compared to patients with an affective disorder. The extent of support of patients with a schizophrenia spectrum disorder was predicted by age, whereas the feeling of self-efficacy predicted data entry pace.

**Conclusion:** Our results indicate that, although patients with a schizophrenia spectrum disorder need more support and more time for data entry than patients with an affective disorder, digital data collection using patients’ self-reports is a feasible and well-received method. Future clinical and research efforts on digitized assessments in psychiatry should include patients with a schizophrenia spectrum disorder and offer adequate support to reduce digital exclusion of these patients.

## Introduction

Schizophrenia spectrum disorders (SSDs) comprise psychiatric diagnoses such as schizophrenia, schizoaffective disorder, schizotypal and delusional disorder which are described in ICD 10 in sections F20.0 through F29.9 (World Health Organization, 1993). Schizophrenia as the most relevant disorder of this spectrum is a frequently chronic psychiatric disorder with a life-time prevalence of almost one percent (Kahn et al., 2015). In spite of a relatively low prevalence it is among the world’s top ten reasons for long-term disability (Mueser & Mcgurk, 2004) which, in addition to its impact on the individual, highlights the large economic and societal burden of this disorder. Successful treatment is difficult due to the considerable heterogeneity of the clinical picture of patients with a schizophrenic spectrum disorder (PSSD). It has long been considered as a syndrome consisting of different subtypes (Ahmed et al., 2018; Buchanan RW, 1994). There is a high susceptibility to relapse in schizophrenia. Forty percent of inpatients suffer from a relapse within one year even though they received appropriate therapy (Barnett et al., 2018) and each new episode increases the risk of chronicity (Alvarez-Jimenez et al., 2012). Hence, it has been suggested that detailed information on risk and symptom profiles might help to identify patients that are particularly vulnerable to relapse (Habtewold et al., 2020). The implementation of data collections during inpatient treatment for schizophrenia could help to quantify symptom development and treatment response. This would allow for the partial decryption of the disorder and identify patients that are at great risk to relapse (Henson et al., 2021; Torous et al., 2018). Ideally, this data is assessed digitally as it can be stored directly in the patient’s electronic medical record and the clinical staff has a direct access to the patients’ information. Many therapeutic processes can be accelerated and practitioners are offered a chance to get a holistic understanding of their patients’ health (Hsin et al., 2018; Mehta et al., 2019; Rojnic Kuzman et al., 2018).

There are plenty of ways to use digital data in psychiatry, still the usage of digital resources is little (Myin-Germeys, 2020) and we are still early in the process of converting from paper to a digital based medicine (Torous & Baker, 2016). Big data approaches in psychiatry face particular challenges, as their implementation relies on the clinicians but also strongly on the participation of patients (Monteith et al., 2016). Certain relevant information on mood, affective state or psychotic symptom severity cannot be assessed externally and must therefore be provided by the patients themselves. Self-reports gain ground, as they enable to actively participate in research efforts (Sartorius, 2014). Presented in a digital format they can be used to collect data in a location- and time-variable manner. Compared to classic paper questionnaires, digital assessment tools are of similarly high reliability (Alfonsson et al., 2014; Hsin et al., 2018). They are more cost-saving (Kuzman et al., 2017; Marcano Belisario et al., 2014) and have been shown to be preferred among medical staff as well as non-psychiatric patients (Fritz et al., 2012). Touchscreen modules in particular are gaining increasing acceptance (Preuschoff et al., 2013). We have already established a patient-reported outcomes system at the University Hospital Münster and showed its validity in patients with an affective disorder (PADs; Richter et al., 2020).

PSSDs however are often not trusted with handling the digital opportunities that come with our time. Concerns exist about the user engagement (Ben-Zeev et al., 2016; Daker-White & Rogers, 2013; Surmann et al., 2017) and that cognitive impairment may complicate the use of assistive technologies (Surmann & Lencer, 2017; Treisman et al., 2016). As a consequence patients find themselves digitally excluded with less use of computers, mobile phones and the internet (Greer et al., 2019; Wong et al., 2020). This led Firth and Torous (2015) to state that PSSDs face a double stigma based on the nature of their condition on the one hand, and attitudes toward their abilities to handle digital media on the other. According to Robotham (2016) there still seems to exist a “digital divide” with the risk that PSSDs benefit less from digitalization than other patient groups. It is suggested that four out of five patients with schizophrenia and related psychoses would not fit criteria to enroll in a typical treatment research study (Humphreys, 2017) demonstrating a considerable selection bias and systematic exclusion of more acutely ill PSSDs from research efforts. The development towards a more digitized psychiatry bears the risk that PSSDs are excluded even more from progresses in research (Kidd et al., 2019; Robotham et al., 2016; Treisman et al., 2016).

Furthermore it could be expected that a paranoia towards digital media lies within the nature of PSSDs (McLaren et al., 1995; Santesteban-Echarri et al., 2020; Treisman et al., 2016). Patients with severe mental health problems seem to be at risk of misinterpreting the virtual world and suffer from paranoia after the use of social media for instance (Berry et al., 2018). However, current research indicates that there seems to be willingness and desire of patients to integrate digital tools in their everyday life (Ben-Zeev et al., 2013; Berry et al., 2019; Bucci et al., 2018).

Yet, only few studies have examined the applicability with PSSDs, whether for complex data collection or digital interventions (Barnett et al., 2018; Kidd et al., 2019; Liu et al., 2019; Tolley et al., 2015). Previous studies are characterized by small samples of psychopathologically stable patients and have typically focused on outpatients. Furthermore, research efforts did not include insights about the patients’ subjective experience with the digital medium. While there are studies exploring the perception of medical staff towards digital data assessments in psychiatry (Aref-Adib et al., 2020; Odendaal et al., 2020), to the best of our knowledge, no validated psychometric instrument exists for the assessment of PSSDs’ subjective experience with digital data collections in clinical routine.

In spite of the existing stigmas, we hypothesize that the digital exclusion of PSSDs is to further extent unjustified. To test this hypothesis, the present controlled feasibility study investigates the applicability of a longitudinal digital data assessment with the help of a tablet computer during inpatient treatment of PSSDs. We investigated the acceptance, the level of support and the time required to perform the data entry. Results are compared to those of PADs. Lastly sociodemographic and psychometric factors on the handling of the tablet of PSSDs are considered.

## Methods

### Sample

A total of 182 inpatients of the Department of Psychiatry, University Hospital Münster, was approached and asked to participate in the study between February 2020 and February 2021. Criteria for inclusion were a diagnosis of schizophrenia, a schizoaffective, a schizotypal, delusional and other non-mood psychotic (F20.0 through F29.9) or affective (F30.0 through F39.9, ICD-10) disorder and sufficient German language knowledge. Please refer to Table S1 and Table S2 for further information on the participants’ diagnoses. The study was approved by the institutional review board of the Medical Faculty, University of Münster, and written informed consent was obtained from every patient. We predefined reasons for exclusion: organizational reasons, exclusion by clinicians because of severe cognitive deficits or mental instability that would hinder participation and insufficient German language knowledge.

### Procedure

We aimed to achieve a number of 100 participating patients and proceeded recruitment until we included 49 PSSDs and 51 PADs. The recruitment steps followed procedures described in our previous work (Richter et al., 2020). Patients with the appropriate diagnosis were identified through a patient recruitment system based on the diagnosis entered into the electronic health record by the attending physician at admission. Patients were informed in detail about the study’s main objective (digital assessment of risk and resilience factors for relapse and chronification) and were invited to answer tablet-based questions every other week during their hospital stay. Patients were also informed that the clinical staff had access to their answers.

The tablet-based baseline assessment included self-report questionnaires on sociodemographic data, personal and family mental health history, the Childhood Trauma Questionnaire (CTQ, Bernstein et al., 1997), the Big Five Inventory (BFI, Soto & John, 2017), the Self-Efficacy Scale (SES, Luszczynska et al., 2005) and Beck’s Depression Inventory (BDI, Beck et al., 1960). The Hamilton Depression Scale (HAMD, Hamilton, 1960), the Global Assessment of Functioning (GAF, Hall, 1995) and the Positive and Negative Syndrome Scale (PANSS, Kay et al., 1987) were additionally carried out by trained research assistants. Patients entered data via the Mobile Patient Survey, a web-based multi-language electronic patient-reported outcome system (Soto-Rey et al., 2017) on an Apple iPad tablet. Completed data entries were added automatically to the electronic health record. The technical infrastructure for data acquisition, storage and export implemented at the Department of Psychiatry and Psychotherapy has been extensively described in our previous work (Blitz et al., 2021). Finally, patients were asked to evaluate their own performance and contentment regarding the handling of the tablet questionnaire (Table 1).

**Table 1:**
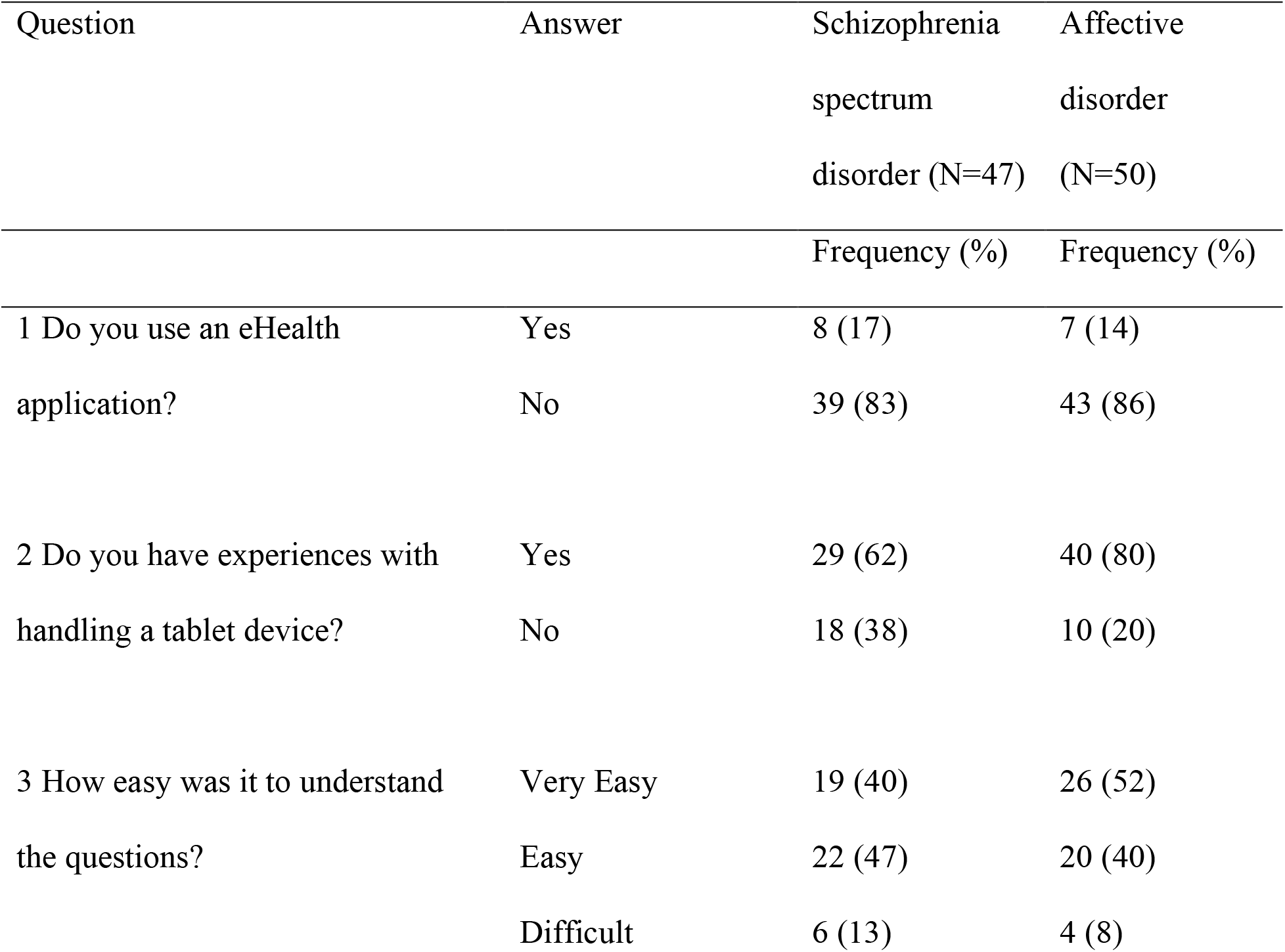
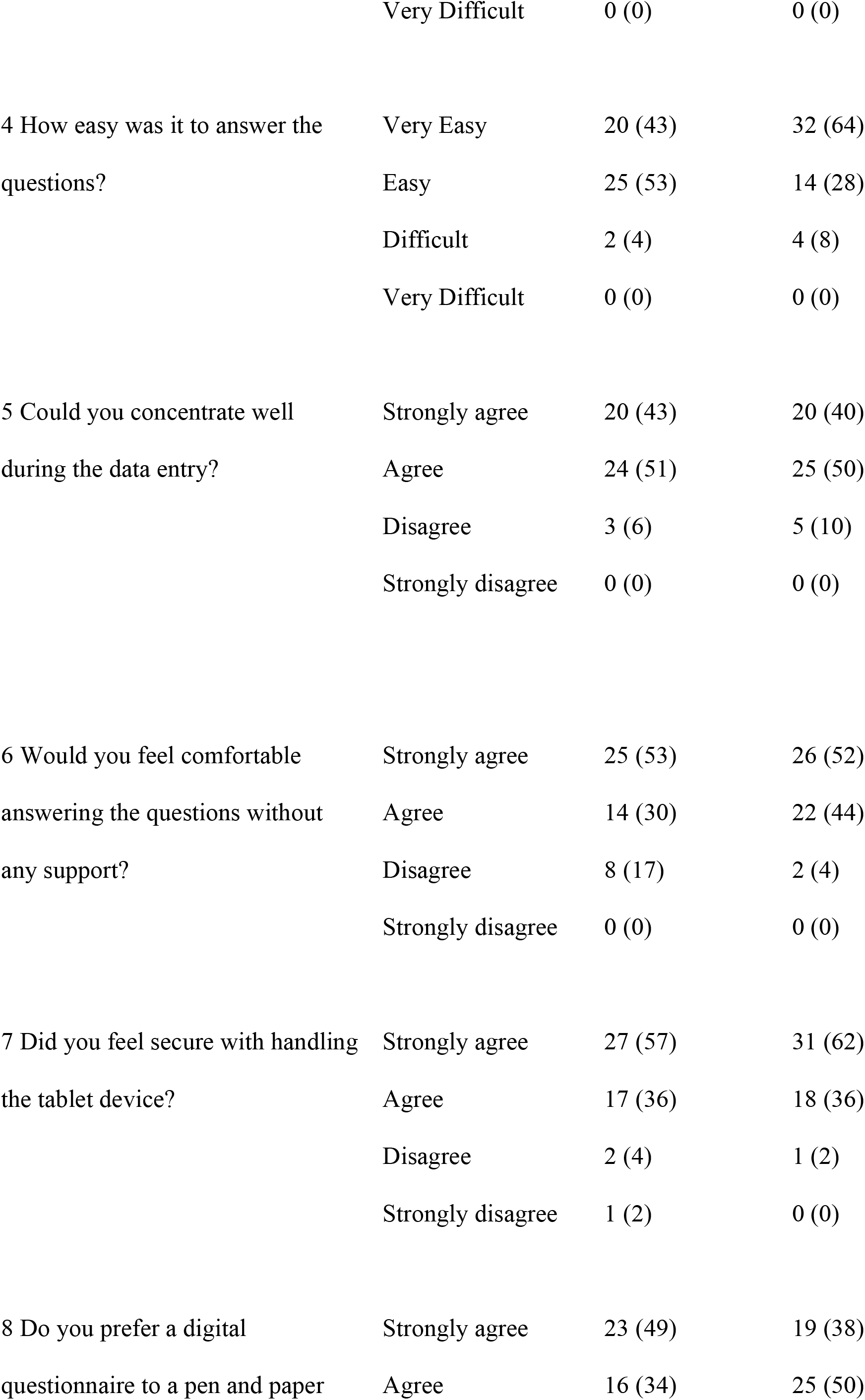
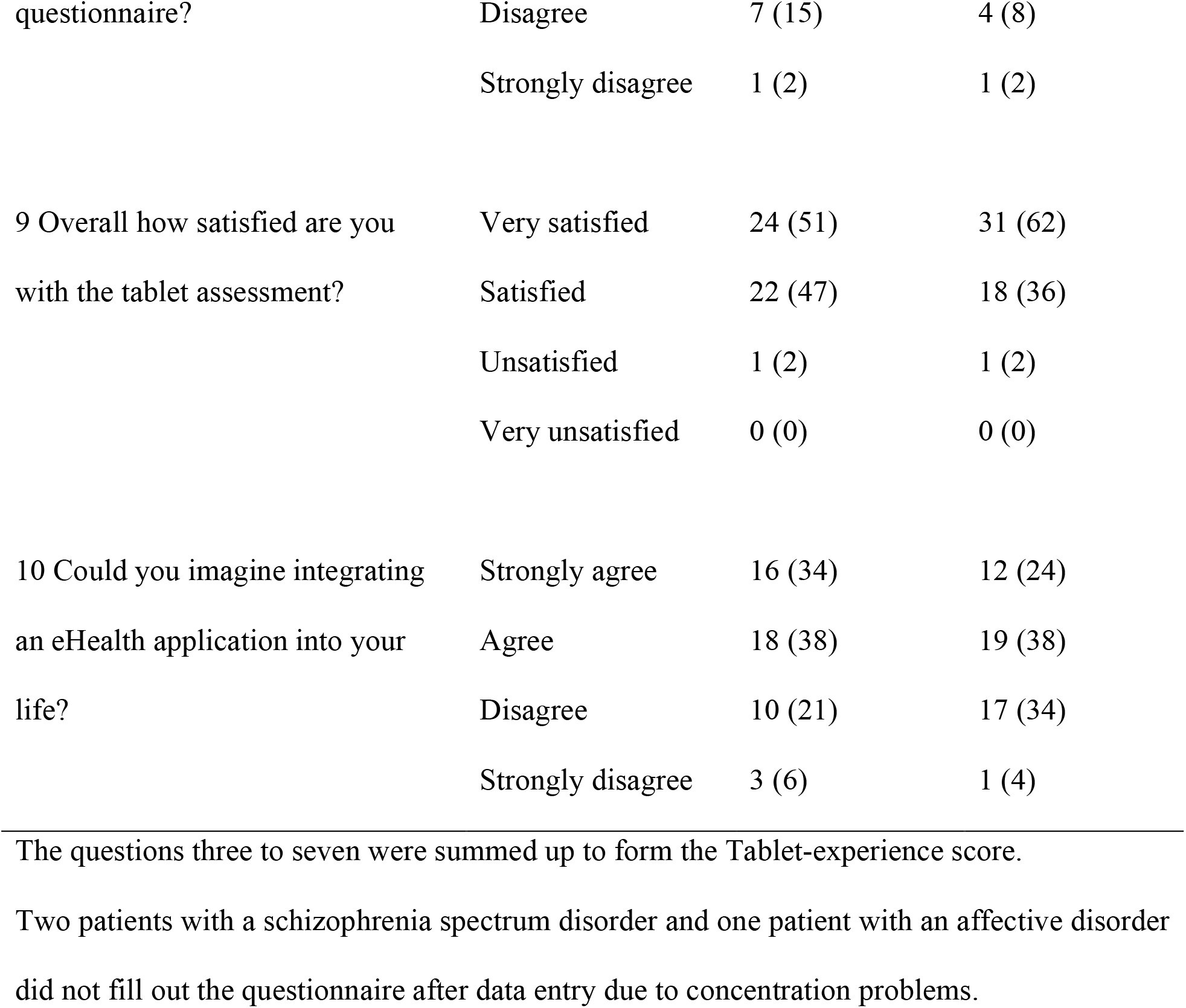
Questionnaire asking about the patient’s subjective experience with the assessment using a tablet

### Acceptance

Due to the absence of a validated psychometric instrument, we developed a novel questionnaire to assess subjective user experience at baseline assessment as well as previous experience with digital health applications. More concretely, to assess patients’ subjective experience of the digital assessment, patients were asked about prior experience with mental health smartphone applications and tablet use, about their individual difficulties understanding and answering the questions on the tablet and whether they were interested in integrating similar questionnaires into their everyday life by means of a smartphone application (Table 1). These data were assessed in form of a paper-pencil questionnaire after data entry.

Based on the questions 3 to 7 (Table 1) and a 4-point Likert-scale (very easy/definitely=4; very difficult/not at all=1) a sum-score (Tablet-experience score) was created with 20 as the highest and 5 as the lowest score. The total score gives an estimate about how well the patient understands and answers the questions. The internal consistency of the score was satisfying with a Cronbach’s alpha of .84.

We performed a linear regression model to assess the influence of age, gender, depressive symptom expression, the level of functioning and self-efficacy on the Tablet-experience score of PSSDs. Age, gender, the baseline sum scores of BDI, HAMD, GAF and SES served as independent variables while the Tablet-experience score served as the dependent variable.

### Support and Data Entry Pace

During assessment a research assistant was present to offer support to patients if required and to record the time of data entry. Additionally, the support with completing the data entry was rated by the research assistant by means of a four-point Likert scale: 1, no support: patient enters data independently; 2, little required support: patient needs few instructions before entering data; 3, some required support: patient needs instructions several times; 4, a lot of support: for the most part the patient is dependent on help. The average support and the mean time for data entry (in minutes) at baseline were assessed.

To assess the relation between age, gender, level of functioning, depressive and psychotic symptoms and required support of PSSDs we estimated an ordinal logistic regression model. Age, gender, baseline sum scores of HAMD, GAF, PANSS and general psychopathology sub score of the PANSS represented the independent variables while support was defined as the dependent variable.

Similarly, we assessed the relation between age, gender, HAMD, GAF and PANSS with data entry pace of PSSDs. The independent variables consisted of age, gender, the baseline sum scores of HAMD, GAF and PANSS and the general psychopathology scale of the PANSS. Data entry pace served as the dependent variable.

### Statistical analyses

Statistics were computed using the SPSS software package (version 26; IBM Corp). We aimed to explore the patients’ acceptance, the performance and the required time to carry out the digital data assessment and had a special interest in the differences between PSSDs and PADs. In order to compare the subjective experience, the required support and data entry pace in the two patient groups we used two-tailed sample t-tests and chi-square tests. We followed the same procedure when comparing the age and gender of participants and nonparticipants. As the data on symptom severity was not available for nonparticipants we could not assess differences in these measures.

Regression analyses were performed in order to identify variables that were significantly associated with the subjective experience, support and data entry pace of PSSDs.

For all models, uncorrected *P* values as well as Benjamini-Hochberg false discovery rate– corrected *P* values are reported.

## Results

### Demographics

100 patients participated in the study of whom 49 (49%) were diagnosed with a schizophrenia spectrum disorder and 51 (51%) with an affective disorder.

Out of 82 approached PSSDs 49 (59.8%) agreed to participate in the study, while 33 (40.2%) patients did not meet inclusion criteria or refused participation (Figure 1). 51 (51%) PADs participated in the study during the outlined recruitment period, 49 (49%) either could not be included or refused participating (Figure 2).

**Figure 1.**
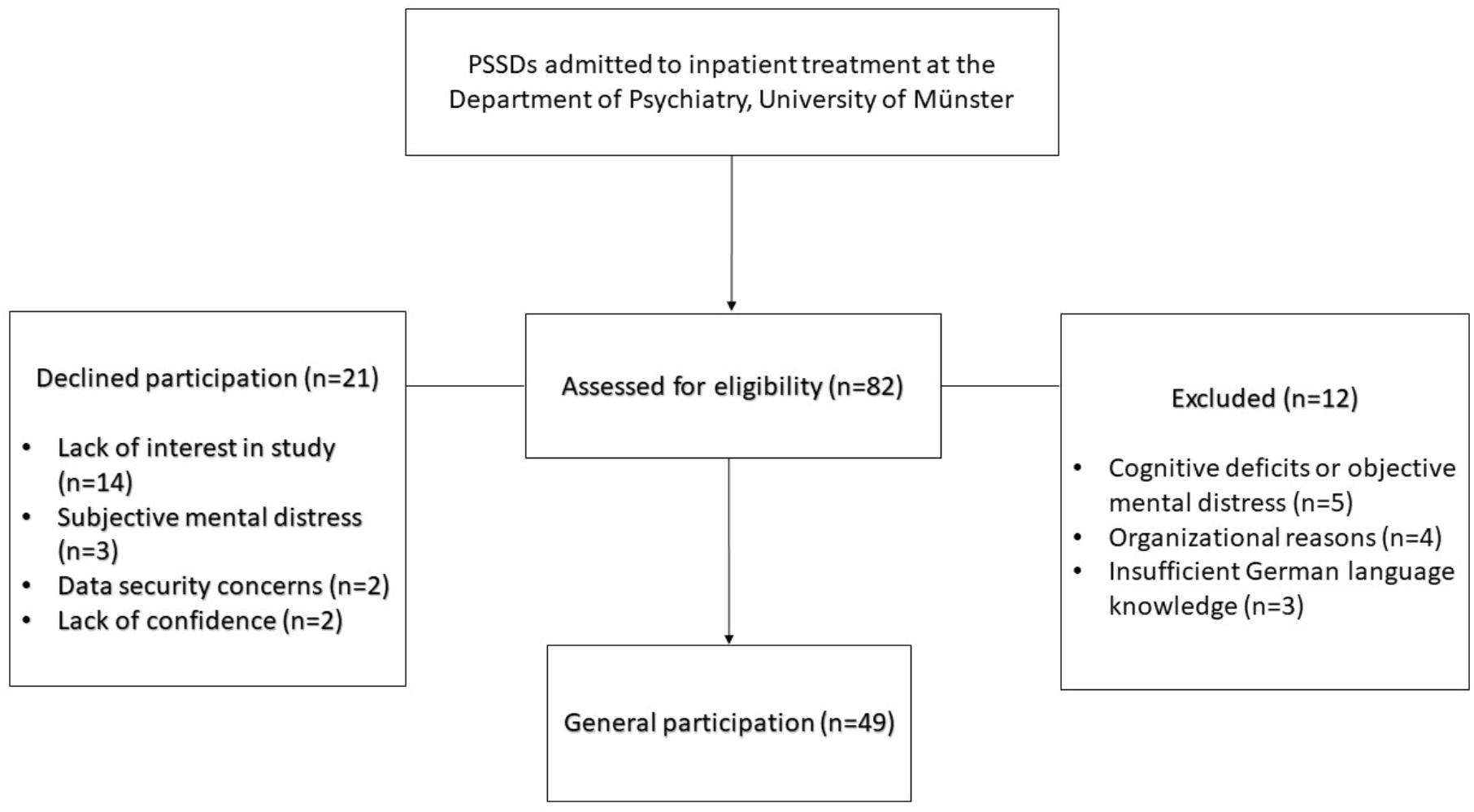
Study flow chart: Patients with a schizophrenia spectrum disorder (PSSDs)

**Figure 2.**
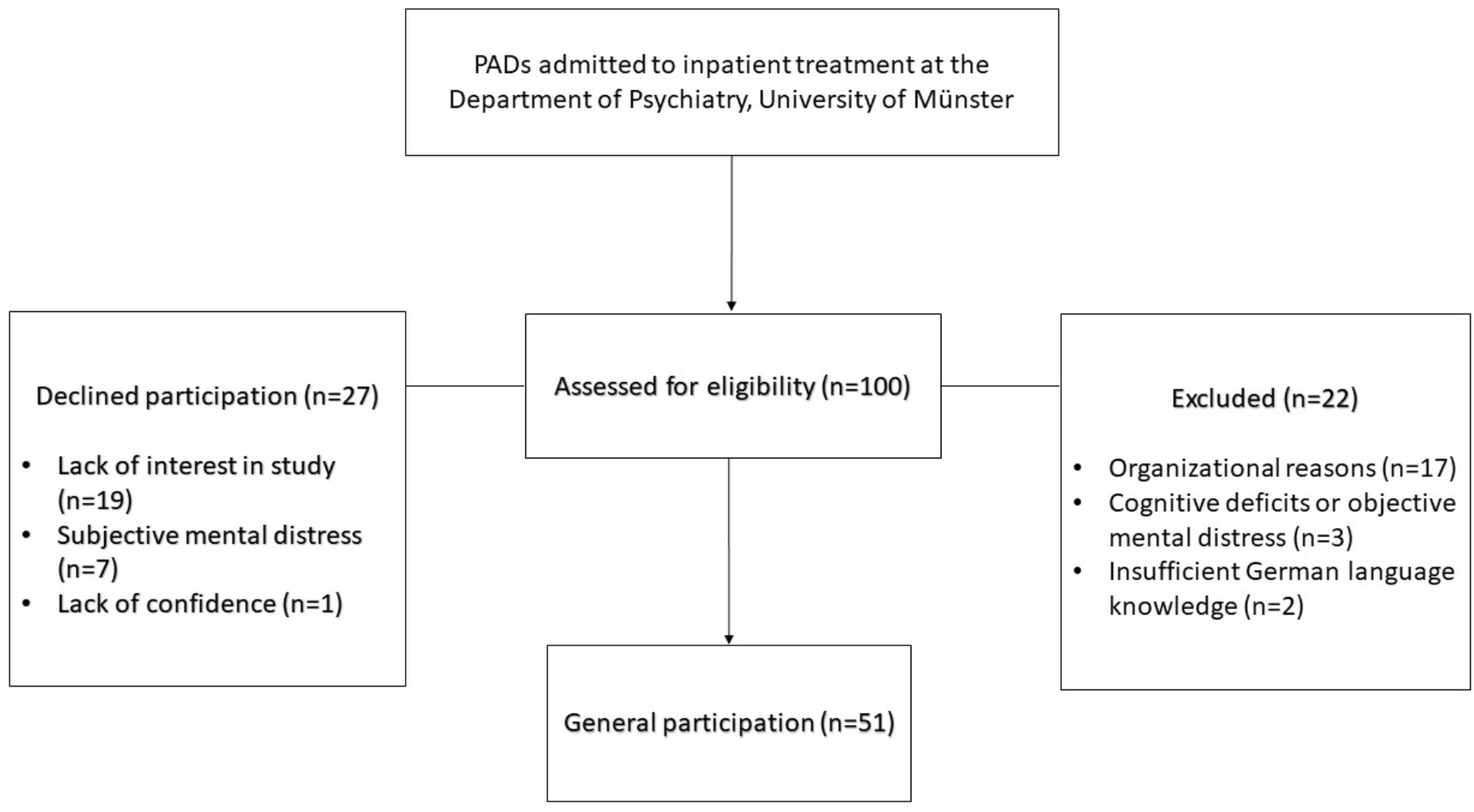
Study flow chart: Patients with an affective disorder (PADs)

We found no statistically significant differences neither in the participation (X^2^=1.395, P=.237) nor the refusal rate (X^2^=.045, P=.832) between both patient groups.

Mean duration of hospitalization of PSSDs was 56.39 (SD=37.48) days with on average 2.79 (SD=1.69) and a median of two assessments (range 1-7). Mean duration of hospitalization of PADs was 52.60 (SD=23.68) days with an average of 3.48 (SD=1.66) and a median of four assessments (range 1-8).

The two patient groups did not show significant differences in age (t_98_=.366, P=.715, P_FDR_=.737.), duration of illness (t_93_=-1.103, P=.273, P_FDR_=.387) and years of education (X^2^=8.63, P=.196, P_FDR_=.340). Gender distribution differed between PSSDs and PADs with higher frequency of men in the SSD group (Table 2)

**Table 2:**
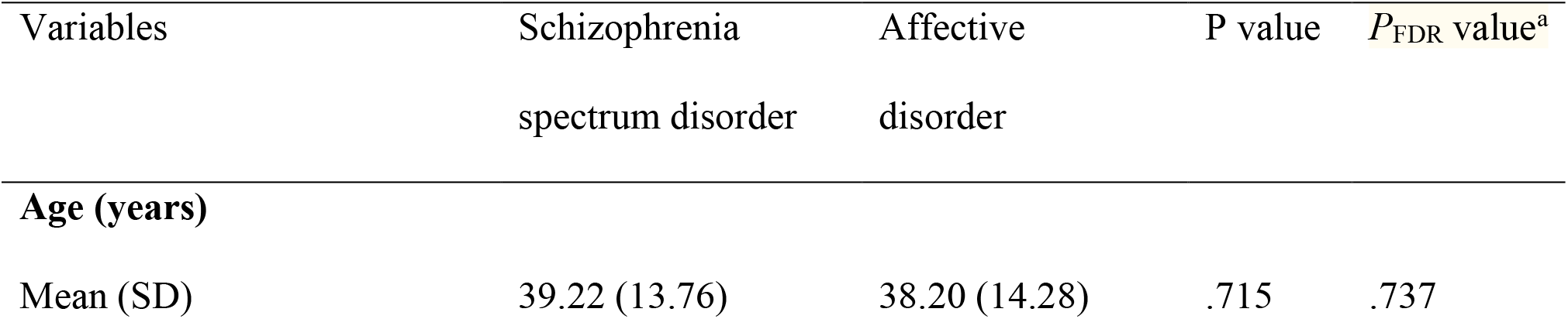
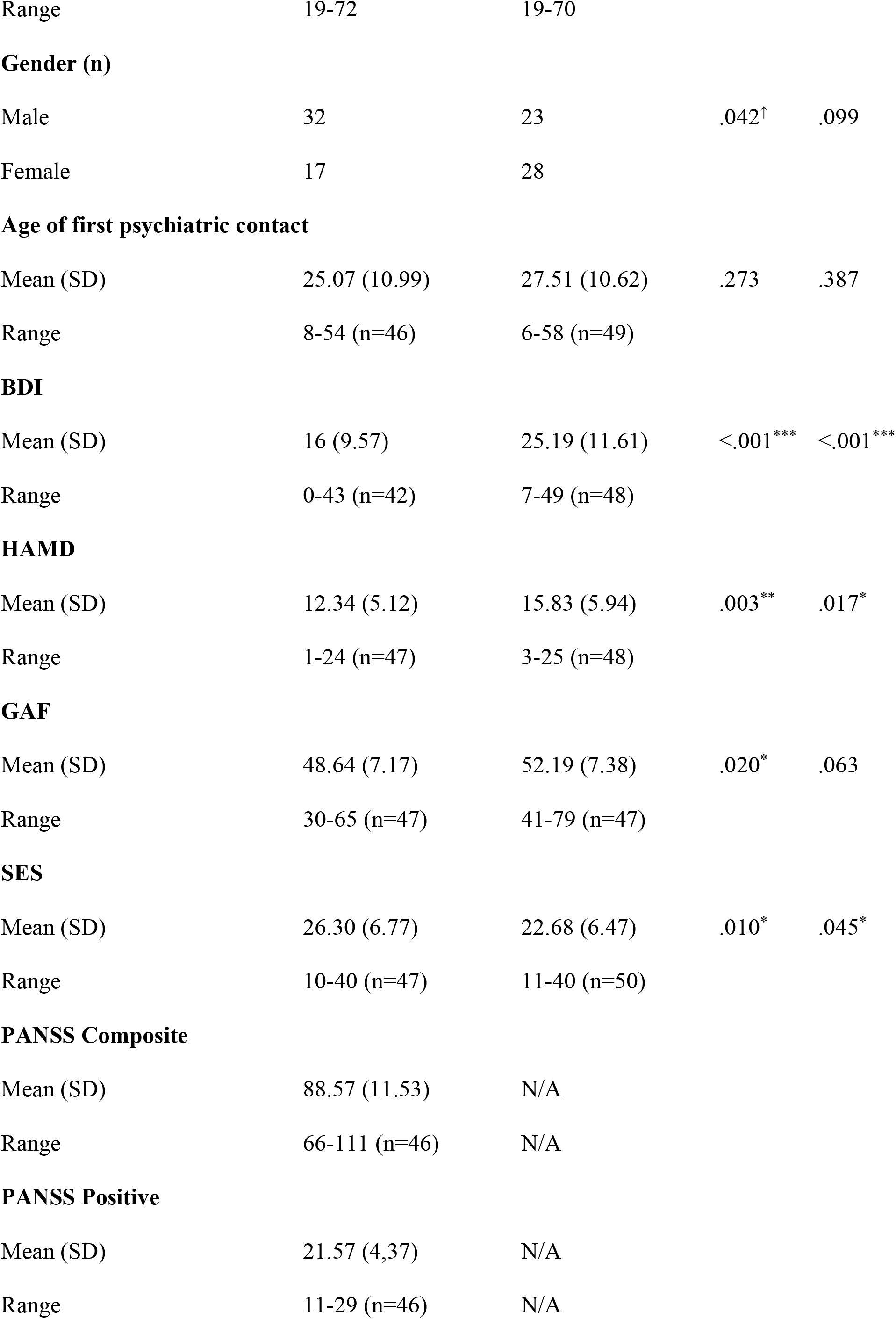
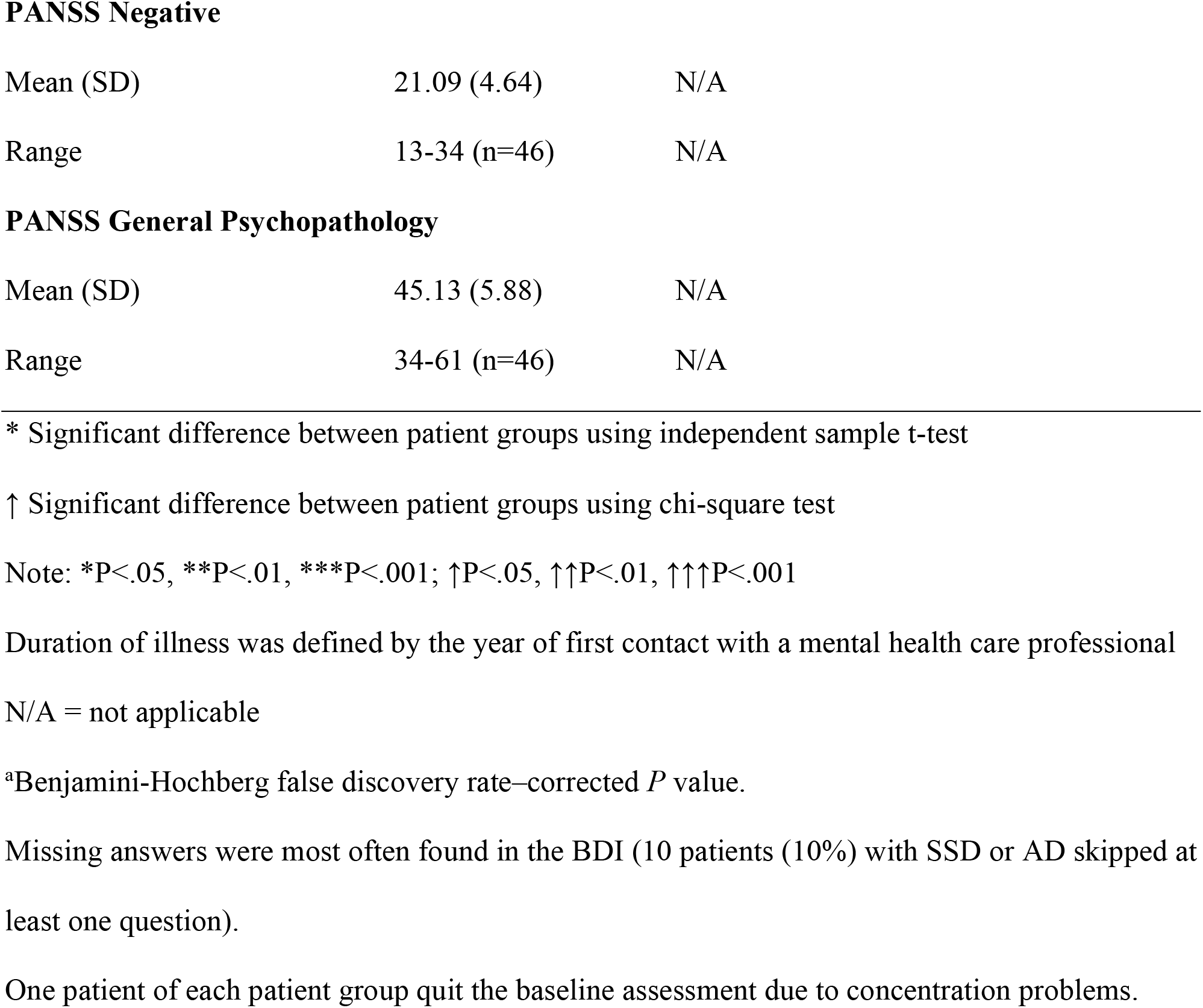
Sociodemographic and clinical characteristics of PSSDs and PADs at baseline assessment

| Variables | Schizophrenia<br>spectrum disorder | Affective<br>disorder | P value | $P_{FDR}$ value<sup>a</sup> |
| --- | --- | --- | --- | --- |
| **Age (years)** |  |  |  |  |
| Mean (SD) | 39.22 (13.76) | 38.20 (14.28) | .715 | .737 |
| Range | 19-72 | 19-70 |  |  |
| **Gender (n)** |  |  |  |  |
| Male | 32 | 23 | .042<sup>†</sup> | .099 |
| Female | 17 | 28 |  |  |
| **Age of first psychiatric contact** |  |  |  |  |
| Mean (SD) | 25.07 (10.99) | 27.51 (10.62) | .273 | .387 |
| Range | 8-54 (n=46) | 6-58 (n=49) |  |  |
| **BDI** |  |  |  |  |
| Mean (SD) | 16 (9.57) | 25.19 (11.61) | <.001<sup>***</sup> | <.001<sup>***</sup> |
| Range | 0-43 (n=42) | 7-49 (n=48) |  |  |
| **HAMD** |  |  |  |  |
| Mean (SD) | 12.34 (5.12) | 15.83 (5.94) | .003<sup>**</sup> | .017<sup>*</sup> |
| Range | 1-24 (n=47) | 3-25 (n=48) |  |  |
| **GAF** |  |  |  |  |
| Mean (SD) | 48.64 (7.17) | 52.19 (7.38) | .020<sup>*</sup> | .063 |
| Range | 30-65 (n=47) | 41-79 (n=47) |  |  |
| **SES** |  |  |  |  |
| Mean (SD) | 26.30 (6.77) | 22.68 (6.47) | .010<sup>*</sup> | .045<sup>*</sup> |
| Range | 10-40 (n=47) | 11-40 (n=50) |  |  |
| **PANSS Composite** |  |  |  |  |
| Mean (SD) | 88.57 (11.53) | N/A |  |  |
| Range | 66-111 (n=46) | N/A |  |  |
| **PANSS Positive** |  |  |  |  |
| Mean (SD) | 21.57 (4.37) | N/A |  |  |
| Range | 11-29 (n=46) | N/A |  |  |

**PANSS Negative**
|  |  |  |
| --- | --- | --- |
| Mean (SD) | 21.09 (4.64) | N/A |
| Range | 13-34 (n=46) | N/A |

**PANSS General Psychopathology**
|  |  |  |
| --- | --- | --- |
| Mean (SD) | 45.13 (5.88) | N/A |
| Range | 34-61 (n=46) | N/A |

\* Significant difference between patient groups using independent sample t-test
↑ Significant difference between patient groups using chi-square test
Note: \* $P < .05$ , \*\* $P < .01$ , \*\*\* $P < .001$ ; ↑ $P < .05$ , ↑↑ $P < .01$ , ↑↑↑ $P < .001$
Duration of illness was defined by the year of first contact with a mental health care professional
N/A = not applicable
<sup>a</sup>Benjamini-Hochberg false discovery rate–corrected $P$ value.
Missing answers were most often found in the BDI (10 patients (10%) with SSD or AD skipped at least one question).
One patient of each patient group quit the baseline assessment due to concentration problems.

The nonparticipating group of PSSDs consisted of more women than the participating group. However, false discovery rate corrected p-values indicated no statistically significant association between gender and participation (n=82, X^2^=5.38, P=.21, P_FDR_=.063). The nonparticipating group of PADs was older than patients of the participating group (t (98) =3.51, P=.001, P_FDR_=.011). There was no difference in age between the participating and nonparticipating group of PSSDs (t (80) =1.06, P=.291, P_FDR_=.387). Please refer to Table S7 for more information on Nonparticipants.

### Acceptance

The assessment was positively received by both subsamples; 98% of patients in both groups were either satisfied or very satisfied with the data entry (Table 1, question 9). The individual Tablet-experience score indicating the patient’s subjective confidence and security during data entry did not differ significantly between the two diagnosis groups (t(95)=-.880, p=.381, P_FDR_=.449; Table 3). 72% of PSSDs and 62% of PADs could imagine integrating an eHealth app into their daily life (Table 1, question 10). We found, that higher Tablet-experience score values correlated with less required support by patients (r(95)=-.498, p<.001) and a faster data entry pace (r(95)=-.430, p<.001; Table S3).

**Table 3:** Group comparison on the Tablet-experience score

| Variable | Schizophrenia<br>spectrum disorder | Affective<br>Disorder | P value | $P_{FDR}$ value<sup>a</sup> |
| --- | --- | --- | --- | --- |
| **Tablet-experience score** |  |  |  |  |
| Mean | 16.87 | 17.32 | .381 | .449 |
| Range | 12-20 (n=47) | 11-20 (n=50) |  |  |

<sup>a</sup>Benjamini-Hochberg false discovery rate-corrected $P$ value.

The variables used in our regressions showed a significant Pearson correlation in a pre-performed correlation matrix (Table S3).

Predictor variables for the Tablet-experience score were tested by means of a linear regression model. We checked for multicollinearity of the independent variables using the tolerance and the variance inflation factor (VIF). Results indicated that multicollinearity was not a concern (Age, Tolerance= .847, VIF=1.180; Gender, Tolerance=.923, VIF= 1.084; HAMD, Tolerance= .494, VIF= 2.023; GAF, Tolerance= .827, VIF= 1.210; SES, Tolerance= .599, VIF= 1.668). The model was significant and explained 45.7% of the variance in the Tablet-experience score (R^2^=.457, F=6.238, P<.001). Age, gender, HAMD, GAF and SES served as independent variables whereas the Tablet-experience score served as the dependent variable. Age and HAMD were found to be statistically significant negative predictors of the model (Table S4). However, associations did not uphold when correcting for multiple comparisons.

### Support and data entry pace

Approximately half of PSSDs (26/54.2%) and the majority of PADs (41/82%) could enter data without any support (Figure 3). PSSDs needed significantly more support compared to PADs (X^2^=11.21, P=.011, P_FDR_=.045; Table 4).

**Figure 3:**
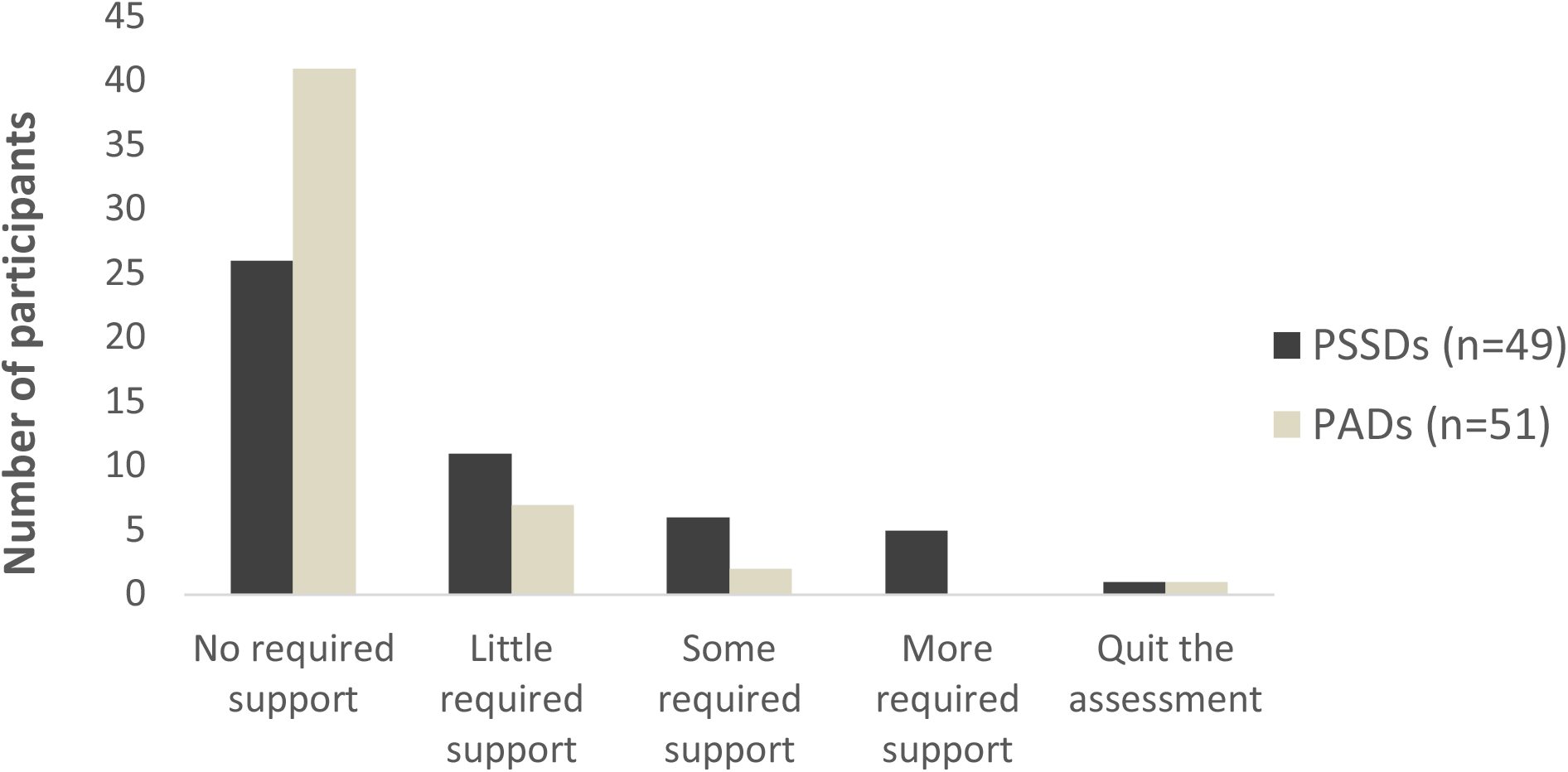
Required support during data entry (n=100)

**Table 4:**
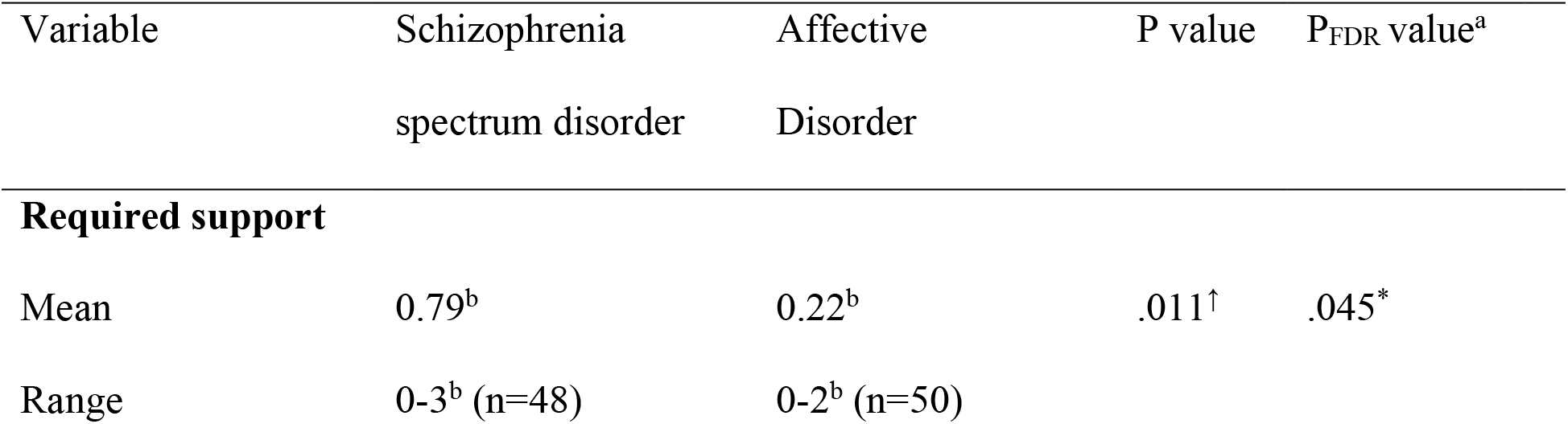
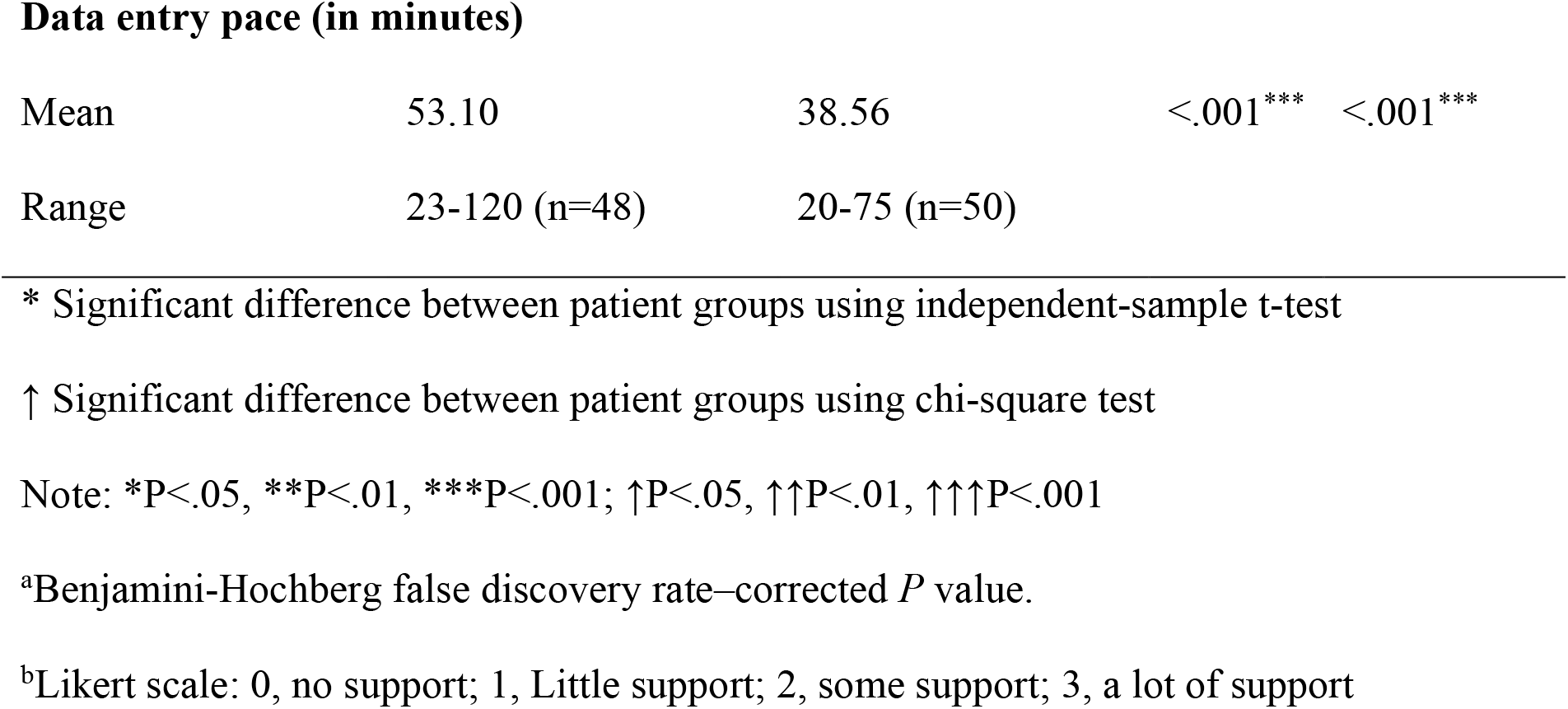
Group comparisons on required support and data entry pace during data entry

| Variable | Schizophrenia<br>spectrum disorder | Affective<br>Disorder | P value | $P_{FDR}$ value<sup>a</sup> |
| --- | --- | --- | --- | --- |
| **Required support** |  |  |  |  |
| Mean | 0.79<sup>b</sup> | 0.22<sup>b</sup> | .011<sup>†</sup> | .045* |
| Range | 0-3<sup>b</sup> (n=48) | 0-2<sup>b</sup> (n=50) |  |  |
| Data entry pace (in minutes) |  |  |  |  |
| Mean | 53.10 | 38.56 | <.001*** | <.001*** |
| Range | 23-120 (n=48) | 20-75 (n=50) |  |  |

\* Significant difference between patient groups using independent-sample t-test
↑ Significant difference between patient groups using chi-square test
Note: \*P<.05, \*\*P<.01, \*\*\*P<.001; ↑P<.05, ↑↑P<.01, ↑↑↑P<.001
<sup>a</sup>Benjamini-Hochberg false discovery rate-corrected *P* value.
<sup>b</sup>Likert scale: 0, no support; 1, Little support; 2, some support; 3, a lot of support

PSSDs needed on average 53.1 minutes (SD=22.94) to complete the assessment at baseline while PADs needed 38.56 minutes. There was a statistically significant difference in time (t(96)=3.86, P<.001, P_FDR_<.001; Table 4).

Predictors for required support of PSSDs were tested by means of an ordinal logistic regression model. The model fit of the regression was given, X^2^=28.930, P= .001. According to Nagelkerke *R*^2^, the model explained 53.2% of the variance. Age, gender, HAMD, GAF, PANSS composite and PANSS general psychopathology served as independent variables and the level of required support as the dependent variable. Age was found to be the only significant predictor of required support. The odds of needing assistance for the data entry increased with age (odds ratio [OR] 1.10, 95% CI 1.03-1.17; P=<.003, P_FDR_=.017; Table S5).

The linear regression model for the data entry pace of PSSDs was significant and explained 36.9% of the variance in data entry time (adjusted R^2^=.369, F=5.001, P=.001). The tolerance and VIF indicated that multicollinearity was not a concern (Age, Tolerance= .81, VIF=1.23; Gender, Tolerance= .76, VIF= 1.33; HAMD, Tolerance= .53, VIF= 1.89; PANSS, Tolerance= .17, VIF= 5.82; General symptom psychopathology scale, Tolerance= .17, VIF= 5.78; SES; Tolerance= .61, VIF= 1.65). Age, gender, HAMD, PANSS composite, PANSS general psychopathology and the SES were the independent variables and the time in minutes for data entry the dependent variable. The SES, the PANSS and the age of patients contributed significantly to the model. The SES was found to be a statistically negative predictor, whereas the PANSS and age contributed positively to the model (Table S6). The associations of the PANSS and age did not uphold when testing for multiple comparisons.

## Discussion

We investigated the acceptance and feasibility of a digital data collection routine during inpatient psychiatric treatment based on self-reports in PSSDs in direct comparison to PADs. Our main findings indicate that PSSDs were equally motivated and willing to engage in digital data collection compared to PADs but needed more support and time for the completion of digital assessments.

While previous studies already showed the feasibility of digital data assessments in outpatient treatment of PSSDs (Liu et al., 2019; Tolley et al., 2015) this study is, to our knowledge, the first demonstrating the acceptance and feasibility of this approach in inpatients in a naturalistic clinical environment. Of all patients who met the inclusion criteria 59.8% of PSSDs and 51% of PADs agreed to participate in this study. These percentages are comparable to studies in the general population (Grobbee et al., 2005) or in non-psychiatric cardiovascular patients (Asselbergs et al., 2017). The diagnosis of a disorder in the spectrum of schizophrenia does not seem to negatively affect the participation rate in a digital data collection effort in clinical routine.

Gathering digital data was assumed to aggravate paranoid ideas in PSSDs, increasing fears of having questionnaire responses tracked and used against them (Chivilgina et al., 2021; Lal et al., 2020). Both PSSDs and PADs showed similar participation rates and gave similar reasons if they refused to participate, which indicates that paranoid ideas did not constitute an obstacle to participate in digitally based data collections, at least in our sample. Only two patients refused to participate because of explicit data security concerns. Few patients declined participation or were excluded by clinicians due to cognitive deficits or symptom severity before being approached. The most common reason for nonparticipation was a general disinterest in the study, which is also one of the main factors for non-use of digital-reported outcome concepts in non-psychiatric patients (Nielsen et al., 2020).

PSSDs were as confident during data entry as PADs and had the impression they could understand and answer questionnaires equally well. This is a very promising result, as previous investigations have demonstrated that PSSDs, in particular older patients, lack confidence in using a computer or a smartphone (Too et al., 2020; Wong et al., 2020). It should be considered that we asked for the patients’ self-evaluation after they entered data with the tablet. It is possible that the patients would have had lower confidence with digital devices if they had been asked before the assessment, as the first assessment could have had a positive effect on their confidence already. Previous data supports that hypothesis and suggests that the use of technology itself leads to self-esteem enhancement (Pourrazavi et al., 2020; Vaportzis et al., 2018). Age had indeed a negative effect on the Tablet-experience score. Even if this trend was not upheld when testing for multiple comparisons, this finding draws attention to not just a psychiatric but societal problem: older adults are still less likely to develop the confidence to engage with digital media and the internet (Gordon & Hornbrook, 2018; Mannheim et al., 2019).

Thirty-eight percent of PSSDs had never used a tablet before being approached for the study and still 98% were either satisfied or very satisfied with the assessment after data entry. Furthermore 72% of patients were open towards installing an eHealth application on their smartphone compared to 62% of PADs, which goes hand in hand with previous studies stating that PSSDs want to get more in touch with digital tools (Ben-Zeev et al., 2018).

Despite the comparatively high participation rate and the overall satisfaction with the assessment, PSSDs had a lower adherence rate than PADs, which can be seen in the mean and median of conducted assessments. When being asked to give an update on their symptom severity every other week fewer PSSDs than PADs were willing to do so. Previous studies have already witnessed lower rates of adherence in new technology based interventions with PSSDs (Alvarez-Jimenez et al., 2014). Killikelly (2017) found male gender and younger age to be specific predictors of nonadherence in mobile and web-based interventions which comprises a big part of the participants with a schizophrenia spectrum disorder in our study. Future studies should keep this in mind and adjust the frequency of assessments to the capabilities and demands of patients.

Regarding the required support, more than half of the PSSDs could read and answer the tablet-based questionnaires independently. Only four patients required a lot of support and relied on the research assistant for the assessment. Overall, PSSDs needed more support and more time for data entry compared to PADs. However, only one patient quit the assessment due to concentration problems and did not finish the data entry at baseline. In conclusion, the diagnosis of a disorder in the schizophrenic spectrum does not seem to have an effect on the dropout rate, a patient might just need to be presented more assistance and patience while handling a digital device. We found the required support to be associated with patients’ age which is in line with studies on general non-clinical samples (Wildenbos et al., 2019) and PADs (Richter et al., 2020). As older adults in general are not as experienced in the use of digital devices this result does not come as a surprise (Mitzner et al., 2019).

The feeling of self-efficacy showed a negative association with the data entry pace of PSSDs. Studies have already shown that low self-efficacy reduces the ability of using a tablet device in older adults (Alvseike & Brønnick, 2012), whereas higher self-efficacy seems to lead to a decrease in computer anxiety (Pourrazavi et al., 2020). There are strategies for increasing the feeling of self-efficacy (Bryce et al., 2018). Clinicians should put a focus on strengthening the patients’ feeling of self-efficacy in order to successfully introduce digital data efforts.

There was a trend for symptom severity (PANSS) to be positively related to the degree of required support. Although this trend was not upheld when testing for multiple comparisons the result is worth discussing. As the PANSS contains aspects like conceptual disorganization, difficulty in abstract thinking, disorientation and poor attention (Kay et al., 1987) it is plausible that the score is associated with the tablet handling performance. However, a greater severity of symptoms was not associated with a higher dropout rate.

While investigation of the validity of patients’ self-reports was not the primary aim of the present study, the strong correlation between the BDI and the HAMD indicated a high validity of patient-reported outcomes. The level of agreement was comparable to what is suggested in the literature (Steer et al., 1987). This is a promising result for the implementation of longitudinal digital data collection for the clinical daily routine and research efforts, as it suggests that no additional clinical personnel is required to gain valid information from PSSDs.

There are limitations to this study. We used the baseline sum scores of BDI, HAMD and SES for our calculations. They could only be applied if all aspects of the questionnaire were answered. Patients were asked to answer all the questions, however they had the opportunity to skip questions. If one question of the instruments above was not answered the total sum score could not be evaluated. We did not assess whether patients skipped questions because they did not understand their meaning or because they did not feel comfortable answering them. However, the great majority of patients answered all questions. The validity of self-assessments was only tested for depression questionnaires. Subsequent studies should also look at the validity of scores that explore the symptoms of a schizophrenia spectrum disorder. Moreover, it should be addressed that our sample size is not big enough to represent the whole clinical population that suffers from schizophrenia spectrum disorders. As this study is one of the first of its kind the sample size is too small to make representative statements. Therefore, future studies should be based on more representative cohorts.

This study explored the feasibility of an implementation of a digital data collection through inpatient treatment of patients suffering from a schizophrenia spectrum disorder in a naturalistic clinical environment. To sum up, while PSSDs needed more support and time for the completion of digital assessments, they were equally open towards them and willing to engage in such efforts compared to PADs. Our findings suggest that digitally assessed self-report measures are well-received in PSSDs and PADs alike and that patients are willing to enter data and give feedback on their symptom severity. As PSSDs were as open towards these technological approaches and showed great confidence while using digital devices, they should be further integrated in digital research efforts. The present results should urge future clinical and research efforts to include PSSDs in digital assessment routines but to account for the higher level of required time and support during digital assessments in PSSDs e.g. by offering personal assistance and ensuring adequate setting during assessments. Achieving these steps could reduce the digital exclusion of PSSDs and hence challenge prevailing stigmas related to SSDs.

## Data Availability

The data that support the findings of this study are available on request from the corresponding author.

## Abbreviations

AD: Affective disorder
BDI: Beck’s Depression Inventory
BFI: Big Five Inventory
CTQ: Childhood Trauma Questionnaire
GAF: Global Assessment of Functioning
HAMD: Hamilton Depression Scale
PADs: Patients with an affective disorder
PANSS: Positive and Negative Symptom Scale
PSSDs: Patients with a schizophrenia spectrum disorder
SES: Self-Efficacy Scale
SSD: Schizophrenia spectrum disorder

## Acknowledgements

We are deeply indebted to all participants of this study. Funding was provided by the Interdisciplinary Center for Clinical Research (IZKF) of the medical faculty of Münster (Grant SEED 11/19 to NO), as well as the “Innovative Medizinische Forschung” (IMF) of the medical faculty of Münster (Grants OP121710 to NO). The study was further supported by a grant from BMBF (HiGHmed 01ZZ1802V).

## Notes

### Competing Interest Statement

The authors have declared no competing interest.

### Author Declarations

The study was approved by the institutional review board of the Medical Faculty, University of Muenster, and written informed consent was obtained from every patient.

