## Supplementary Material for "Symptom Monitoring based on Digital Data Collection During Inpatient Treatment of Schizophrenia Spectrum Disorders – a Feasibility Study"

*Supplementary result*

Depression severity based on patients' self-report (BDI) significantly correlated ($r= .615$, $P<.001$) with an external assessment (HAMD) in PSSDs indicating high validity of digital self-reports (Table S3).

**Table S1:** ICD 10 diagnose of PSSDs

| | Frequency | Percentage |
|---|---|---|
| Diagnose | | |
| F20 Paranoid schizophrenia | 33 | 67.3 |
| F25 Schizoaffective disorder | 10 | 20.4 |
| F23 Polymorphic psychotic episode | 5 | 10.2 |
| F22 Persistent delusional disorder | 1 | 2 |
| Additional psychiatric diagnose | 9 | 11.4 |

**Table S2:** ICD 10 diagnose of PADs

| | Frequency | Percentage |
|---|---|---|
| Diagnose | | |
| F33 Recurrent depressive disorder | 33 | 64.7 |
| F32 Depressive episode | 14 | 27.5 |
| F31 Bipolar affective disorder | 4 | 7.8 |
| Additional psychiatric diagnose | 24 | 47 |

**Table S3:** Summary of intercorrelations between clinical characteristics and performance variables of PSSDs

| | 1 | 2 | 3 | 4 | 5 | 6 | 7 | 8 | 9 | 10 | 11 | 12 | 13 |
|---|---|---|---|---|---|---|---|---|---|---|---|---|---|
| **1 Tablet-experience score** | - | | | | | | | | | | | | |
| **2 Required support** | -.556$^{***}$ | - | | | | | | | | | | | |
| **3 Data entry pace** | -.522$^{***}$ | .564$^{***}$ | - | | | | | | | | | | |
| **4 Age** | -.366$^{*}$ | .620$^{***}$ | .242 | - | | | | | | | | | |
| **5 Gender** | -.204 | .194 | .129 | .279 | - | | | | | | | | |
| **6 HAMD** | -.506$^{***}$ | .418$^{**}$ | .311$^{*}$ | .388$^{**}$ | .151 | - | | | | | | | |
| **7 BDI** | -.478$^{**}$ | .081 | .132 | .028 | .047 | .615$^{***}$ | - | | | | | | |
| **8 GAF** | .328$^{*}$ | -.357$^{*}$ | -.216 | -.206 | -.136 | -.375$^{**}$ | -.185 | - | | | | | |
| **9 PANSS Composite** | -.155 | .403$^{**}$ | .344$^{*}$ | .158 | -.252 | .276 | .202 | -.445$^{**}$ | - | | | | |
| **10 PANSS Positive** | .050 | .219 | .095 | .066 | -254 | .193 | .186 | -.243 | .573$^{***}$ | - | | | |
| **11 PANSS Negative** | -.177 | .264 | .288 | .141 | -.133 | .073 | .088 | -.348$^{*}$ | .648$^{***}$ | -.091 | - | | |

| | | | | | | | | | | | | | |
|---|---|---|---|---|---|---|---|---|---|---|---|---|---|
| **12 PANSS General Psychopathology** | -.255 | .404** | .344* | .179 | -.119 | .378* | .236 | -.480** | .895*** | .348* | .604*** | - | |
| **13 SES** | .414** | -.230 | -.322* | -.084 | .057 | -.615*** | -.586*** | .147 | -.034 | .108 | -.051 | -.154 | - |

Note: *P<.05, **P<.01, ***P<.001

**Table S4:** Linear regression model predicting the effect of age, gender, HAMD, GAF and SES on the Tablet-experience score of PSSDs

| Variables[a] | B (SE) | β | t value | P value | $P_{FDR}$ value[b] |
|---|---|---|---|---|---|
| Age | -0.055 (0.024) | -0.301 | -2.289 | .028* | .074 |
| Gender | -0.708 (0.656) | -0.136 | -1.078 | .288 | .387 |
| HAMD | -0.174 (0.086) | -0.349 | -2.023 | .050 | .103 |
| GAF | 0.018 (0.045) | 0.055 | 0.414 | .681 | .725 |
| SES | 0.060 (0.056) | 0.167 | 1.067 | .293 | .387 |

Note: *P<.05, **P<.01, ***P<.001

[a]Model $R^2$=0.410

**Table S5:** Ordinal logistic regression model predicting the effect of age, gender, HAMD, GAF and PANSS on the required support for data entry of PSSDs

| Variables[a] | B (SE) | Wald | Odds Ratio (95% CI) | P value | $P_{\mathrm{FDR}}$ value[b] |
|---|---|---|---|---|---|
| Age | 0.10 (0.03) | 8.81 | 1.10 (1.03-1.17) | .003** | .017* |
| Gender (male=1) | 1.25 (0.81) | 2.36 | 3.47 (0.71-16.99) | .124 | .227 |
| HAMD | 0.08 (0.08) | 0.93 | 1.08 (0.93-1.26) | .336 | .427 |
| GAF | -0.07 (0.62) | 1.20 | 0.94 (0.83-1.06) | .273 | .387 |
| PANSS Composite | 0.07 (0.79) | 0.80 | 1.07 (0.92-1.25) | .373 | .449 |
| PANSS General Psychopathology | -0.02 (0.15) | 0.02 | 0.98 (0.73-1.32) | .897 | .897 |

Note: *P<.05, **P<.01, ***P<.001

[a]Model Nagelkerke $R^2$=0.532

[b]Benjamini-Hochberg false discovery rate–corrected *P* value

**Table S6:** Linear regression model predicting the effect of age, gender, HAMD, PANSS and SES on the data entry pace for PSSDs

| Variables[a] | B (SE) | β | t value | P value | $P_{FDR}$ value[b] |
|---|---|---|---|---|---|
| Age | 0.54 (0.24) | 0.31 | 2.28 | .029* | .074 |
| Gender | 10.71 (6.49) | 0.24 | 1.65 | .108 | .210 |
| HAMD | -0.64 (0.77) | -0.14 | -0.84 | .408 | .464 |
| PANSS Composite | 1.46 (0.58) | 0.76 | 2.54 | .016* | .059 |
| PANSS General Psychopathology | -1.35 (1.09) | -0.37 | -1.23 | .226 | .373 |
| SES | -1.69 (0.53) | -0.50 | -3.16 | .003** | .017* |

Note: *P<.05, **P<.01, ***P<.001

[a]Model $R^2$=0.217

[b]Benjamini-Hochberg false discovery rate–corrected *P* value.

**Table S7:** Characteristics of participating and nonparticipating patients with a schizophrenia spectrum disorder and an affective disorder

| | Schizophrenia spectrum disorder | | | | Affective disorder | | | |
|---|---|---|---|---|---|---|---|---|
| Variables | Nonparticipants (n=32) | Participants (n=49) | P value | $P_{FDR}$ value | Nonparticipants (n=49) | Participants (n=51) | P value | $P_{FDR}$ value[b] |
| **Age (years)** | | | | | | | | |
| Mean (SD) | 42.48 (13.42) | 39.22 (13.76) | .291 | .387 | 48.82 (15.96) | 38,20 (14.28) | .001** | .011* |
| Range | 20-71 | 19-72 | | | 19-82 | 19-70 | | |
| **Gender (n)** | | | | | | | | |
| Male | 13 | 32 | .021↑ | .063 | 19 | 23 | .527 | .580 |
| Female | 20 | 17 | | | 30 | 28 | | |

Note: *P<.05, **P<.01, ***P<.001; ↑P<.05, ↑↑P<.01, ↑↑↑P<.001

* Significant difference between patient groups using independent-sample t-test

↑ Significance between patient groups using chi-square test

[b]Benjamini-Hochberg false discovery rate–corrected $P$ value
